# Low-risk Features of Emergency Department Patients on a Sepsis Pathway: A Retrospective Cohort Study

**DOI:** 10.64898/2026.10.21.26366119

**Authors:** Benjamin Wyler, Chen Yang, Samuel E. Sondheim, Michael Lanigan, Alejandro Serrato-Guillen, Nathan I. Shapiro, Madhu Mazumdar, Roland C. Merchant

## Abstract

**Introduction:** For adult emergency department (ED) patients initiated on a sepsis care pathway, we sought to identify patient factors associated with favorable clinical and operational outcomes that could help identify patients potentially eligible for sepsis care de-escalation.

**Methods:** We conducted a retrospective cohort study using an electronic health record (EHR) data query from five hospitals in New York City (2022-2024). Multivariable logistic regression models were used to evaluate association of patient demographics, clinical characteristics, and ED diagnoses with four favorable reference outcomes: shorter hospital length of stay (HLOS), no intensive care unit (ICU) stay, survival to discharge, and no sepsis adjudication according to institutional quality-of-care standards.

**Results:** Among 9,443 ED visits, 45.9% of participants were <70 years old; 68.1% were normothermic and 64.7% normotensive; 32.0% had a normal white blood cell (WBC) count; and 17.6% had a normal lactate level. Fewer than one-quarter received an ED diagnosis of sepsis. Overall, 84.5% survived to discharge, 65.9% avoided an ICU stay, 8.5% had an HLOS <72 hours, and 28.0% were not adjudicated as having sepsis. Factors associated with favorable outcomes varied across the four reference outcomes, with younger age, temperature >36C, absence of hypotension, WBC count 4,000-15,000/uL, and lactate <2.0mmol/L linked to multiple outcomes. Age 18-49 years had the strongest association for HLOS <72 hours (adjusted odds ratio (aOR) 3.48, p<0.01) and survival to discharge (aOR 3.15, p<0.01); lack of severe diagnosis had the strongest association for no ICU stay (aOR 2.74, p<0.01); and ED diagnosis other than sepsis or infection had the strongest association for no sepsis adjudication (aOR 8.82, p<0.01). WBC count 4,000/uL to 15,000/uL was the only factor associated with all four outcomes (aORs 1.24-1.70). Patients with a primary ED diagnosis of infection other than pneumonia had more favorable outcomes than those diagnosed with sepsis or pneumonia.

**Conclusion:** Several patient factors may help identify lower risk patients who could be targeted for sepsis care de-escalation after a sepsis pathway has been initiated, although the relevant factors differ by outcome. Further research is needed to evaluate the performance of these factors in prospectively identifying patients most likely to experience favorable outcomes.

## INTRODUCTION

Many emergency departments (EDs) in the United States (US) employ electronic screening processes to identify patients who could have sepsis. ED sepsis electronic screening processes frequently are based on the presence of Systemic Inflammatory Response Syndrome (SIRS) indicators (a.k.a., meeting SIRS criteria), which are common in the setting of infection, but are not helpful by themselves in distinguishing patients with sepsis from those with merely an infection. Previous studies have demonstrated that even though SIRS criteria can identify 71% of patients with a suspected infection as septic, the criteria common in in non-infectious conditions (“sepsis mimics”) and only have specificity of 41% for sepsis. ^1–4^ SIRS also has poor predictive value for serious adverse outcomes of organ failure or mortality. ^5–8^

EDs typically link sepsis screening processes to initiation of sepsis care pathways or “bundles” to ensure timely evaluation and treatment and to comply with guidelines from the Surviving Sepsis Campaign, ^9^ and the Severe Sepsis and Septic Shock Management Bundle (SEP-1) core quality measure utilized by the Centers for Medicare and Medicaid Services (CMS). The decision to initiate a sepsis care pathway is often made before diagnostic studies have been undertaken. Once sepsis treatment is initiated, clinicians may be disinclined to de-escalate care due to institutional pressure to adhere to SEP-1 core quality measures and/or concern of medical-legal consequences. The result of these processes leads to over-treatment, resulting in unnecessary healthcare costs from avoidable hospital admissions, increased ED boarding, and patient harms from nosocomial infections and adverse drug reactions. ^10,11^

There is a clear need to better risk-stratify ED patients meeting SIRS criteria to ensure appropriate treatment of patients with sepsis, avoid diagnostic anchoring on sepsis in non-infected patients, and allow de-escalation of treatment in patients with infection who are at low risk for sepsis and adverse clinical outcomes. Previous efforts to identify better tools to risk stratify patients with suspected sepsis have yielded mediocre results, with both clinical risk stratification tools (e.g. National Early Warning Score-2 (NEWS-2), quick Sequential Organ Failure Assessment (qSOFA)) and biomarkers (e.g. CRP, procalcitonin) having only poor to fair discriminatory ability for sepsis, with AUCs of 0.58-0.75 ^1,5,12–15^

A number of relatively new assays (e.g., Intellisep® (Cytovale®, San Francisco, CA), monocyte distribution width (MDW, Beckman Coulter, Brea, CA), and Triverity™ (Inflammatix™, Sunnyvale, CA) offer promise for improving sepsis diagnosis by identifying pathophysiologic changes associated with immune dysregulation and could help to identify patients in a sepsis pathway in whom sepsis care could be de-escalated. However, employing these tests without consideration of a patient’s pre-test probability of sepsis could result in false positive or false negative test results and consequent errors in clinical decision-making. Knowing which patients might be at lower risk of sepsis and its adverse clinical consequences could assist in choosing the appropriate patient population for utilization of these tests.

It is conceivable that patient factors that are identified during the ED evaluation could help to predict which patients on a sepsis pathway are candidates for de-escalation of sepsis care. While Sepsis-3 criteria—organ dysfunction measured by a change in SOFA score and caused by infection—might seem a logical outcome against which to measure their performance, Sepsis-3 is time-consuming to adjudicate retrospectively and suffers from limited interrater agreement due to uncertainty of infection diagnosis, ambiguity in SOFA scoring, and complexity in attributing of organ dysfunction to infection in patients with complex medical conditions. ^16–19^ Previous research examining patient factors has focused on the risk of adverse outcomes, usually death or ICU stay, often among undifferentiated patients in whom there was an initial clinical suspicion of sepsis or infection, without reference to a sepsis care pathway. ^20–23^ In contrast, to predict which patients are candidates for sepsis care de-escalation, the focus instead should be on the likelihood of favorable outcomes among those already on a sepsis care pathway. There is no established reference standard outcome for this purpose, and the most appropriate reference outcome could depend on the focus of de-escalation—whether it be to facilitate discharge, triage patients to less intensive care, avoid medical-legal risk associated with patient death, or avoid SEP-1 compliance fall-outs.

We aimed to address the need for identifying patient factors that are available during the ED encounter, are associated with a number of better clinical and administrative outcomes, and could be candidates for identifying patients on a sepsis pathway whose sepsis care could be de-escalated. We also aimed to investigate whether the choice of reference standard outcome affects which set of factors are isolated. We selected four favorable clinical and operational outcomes (shorter hospital length of stay (HLOS), avoidance of an intensive care unit (ICU) stay, survival to discharge, and not being adjudicated as a sepsis case by the institution’s sepsis committee) as reference standards. Among adult ED patients placed on a sepsis care pathway in the EDs of a large, urban, multi-site academic health system in New York City, we identified and compared the demographic and clinical characteristics and initial ED diagnostic impressions associated with each of these four reference standard outcomes.

## METHODS

### Study Design, Population and Setting

This retrospective cohort study analyzed electronic health records (EHRs) data from adult patients placed on an ED sepsis care pathway between January 1, 2022 and December 31, 2024, across five New York City hospitals (three in Manhattan, one in Brooklyn, one in Queens) in the same healthcare system. The EDs at these hospitals had a combined total of 348,079 visits in 2024. The health system’s Internal Review Board (IRB) approved the study and granted a waiver of informed consent prior to commencement. The Strengthening the Reporting of Observational Studies in Epidemiology (STROBE) guidelines were followed to ensure appropriate reporting of this observational study (see Supplement Figure S1). ^24^

### Study Participants

We included all ED patients who were placed on a sepsis care pathway across the five participating hospitals. In order to focus on ED clinician-directed care, patients for whom the sepsis pathway was initiated more than six hours after ED arrival (mean/median LOS: 21.0/13.7 hours) were excluded, as these activations were more likely initiated by inpatient teams rather than ED team. We also excluded patients with missing data and those with ED dispositions that would either lead to incomplete outcome data (e.g., discharge, premature departure, transfer to another hospital) or were incompatible with our focus on identifying low-risk patients (e.g., death in ED, transfer to the operating room or to another procedural area directly from the ED). Finally, patients with a recorded age ≥110 years were excluded to avoid misclassification, as ages in this range are assigned in the EHR when a patient’s identity cannot be verified.

### Extracted Data for Analysis

We extracted EHR data on patient demographics (age, sex, race), timing variables (ED arrival time and time of sepsis pathway activation), and initial clinical measurements, including initial temperature, blood pressure, white blood cell (WBC) count, and both initial and repeat serum lactate levels. Additional extracted variables included ED disposition, primary ED diagnosis, length of stay (LOS) (ED, inpatient, and total hospital LOS), ICU admission, and inpatient mortality. As part of routine quality-of-care monitoring and reporting processes, the health care system’s institutional sepsis committee adjudicates potential sepsis cases for the presence of severe sepsis or septic shock during hospitalization using Sepsis-2 criteria based on EHR-recorded indications of whether the clinician suspected that an infection was present, and if the patient had a lactate >4.0 mmol/L or signs of organ dysfunction. We extracted the sepsis adjudication data from the committee’s databases for inclusion in our analysis.

### Classification of ED Diagnoses

In order to facilitate the study analyses, the study’s principal investigator prepared a classification system to group primary ED diagnoses, using as a model the schema described by Linden and Melhammar, which identifies 13 foci of infection and classifies the likelihood of infection on a scale of 1-4. ^25^ Our classification system used seven primary categories related to likelihood of sepsis or infection: (1) sepsis (diagnosis including the term “sepsis”); (2) serious infection (infection for which hospitalization and/or surgical or procedural intervention would be deemed necessary, including central nervous system (CNS) infection, intra-abdominal infection, bacteremia, gangrene, neutropenic fever, intrathoracic infection, and orthopedic infection); (3) infection (all other bacterial, fungal, parasitic, or unspecified infections); (4) viral infection (specified viral etiology); (5) organ failure (diagnosis indicating impairment of organ function without reference to etiology); (6) non-infectious (diagnosis specifying a condition not typically associated with infection); and (7) possible infection (diagnosis for which it was not possible to determine if there was an infectious etiology, typically symptom-related). The “infection” category was further classified into seven subcategories of pneumonia; genitourinary (GU) infection; gastrointestinal (GI) infection; skin and soft tissue infection (SSTI); ear, nose and throat (ENT) infection; febrile syndrome; and other or unspecified infection. Six of the 13 foci identified by Linden and Melhammar were classified as “severe infection” and six were classified as infection. We omitted foreign-body associated infection as a category and added categories for febrile syndrome and viral infection. ED primary diagnoses were separately classified as being severe (associated with presumed need for hospitalization or surgery), non-severe (not inherently requiring hospitalization or surgery), or other (unable to determine need for hospitalization or surgery) (See Supplement Table S-1 for examples of diagnoses in each category).

### Data Analysis

Statistical analyses were conducted using SAS® analytical software (SAS Institute, Inc., Cary, NC). We generated descriptive statistics for patient demographics, clinical variables, and diagnostic categories. HLOS was dichotomized as <72 hours versus ≥72 hours to distinguish between “short stay” clinical observations from longer hospitalizations that typically indicate greater clinical severity or complexity and require greater resource utilization.

#### Univariable and Multivariable Modeling

We first developed univariable logistic regression models to identify demographic and clinical characteristics associated with each of the four primary reference standard outcomes: (1) short stay (HLOS <72 hours, (2) standard care (no ICU-level care during hospitalization, (3) survival to discharge, and (4) not adjudicated as sepsis. Variables were advanced to multivariable logistic regression models if they met any of the following statistical thresholds: p-value <.20 across at least three outcomes, p-value<.05 for at least two outcomes, p-value<.01 for at least one outcome. ^26^ Sex was retained in the multivariable model despite not meeting these criteria because prior studies have demonstrated its association with sepsis outcomes. ^27,28^ Predictors were categorized or dichotomized using established clinical cutoffs and to ensure adequate subgroup sizes for model stability.

## RESULTS

### Participant Sample, Demographics, Clinical Characteristics, and ED Diagnoses

Of the 10,767 ED patients initiated on the sepsis care pathway across the five study hospitals, 1,324 (12.3%) were excluded: 386 (3.6%) who entered the pathway more than six hours after ED arrival; 864 (8.0%) with dispositions of expired (0.6%), discharged (3.2%), premature departure (0.6%), transfer (3.6%), or missing data (0.07%); and 67 (0.6%) with a recorded age ≥110 years. The remaining 9,443 participants are described in Table 1.

**Table 1:** Demographic and Clinical Characteristics of Patients Placed on the Sepsis Pathway (n=9443) Table 1 describes the demographic and clinical features of a cohort of 9,443 patients placed on a sepsis pathway in the emergency department of five hospitals in a major urban health system from 2022-2024, obtained from an electronic health record query

| Age (years) | N | % | Initial WBC count | N | % |
| --- | --- | --- | --- | --- | --- |
| 18-29 | 225 | 2.4% | 4.0-10.9K | 3022 | 32.0% |
| 30-39 | 447 | 4.7% | <4.0K | 681 | 7.2% |
| 40-49 | 627 | 6.6% | 11.0-14.9K | 2088 | 22.1% |
| 50-59 | 1098 | 11.6% | 15.0-19.9K | 1689 | 17.9% |
| 60-69 | 1939 | 20.5% | ≥20.0K | 1963 | 20.8% |
| 70-79 | 2261 | 23.9% | Initial Lactate |  |  |
| 80-89 | 1951 | 20.7% | <2.0 | 1666 | 17.6% |
| 90-99 | 895 | 9.5% | 2.0-3.9 | 4953 | 52.5% |
| Sex |  |  | ≥4.0 | 2696 | 28.6% |
| Male | 5080 | 53.8% | No data | 127 | 1.3% |
| Female | 4361 | 46.2% | Repeat Lactate |  |  |
| Unknown | 2 | 0.0% | <2.0 | 1850 | 19.6% |
| Race/Hispanic ethnicity |  |  | 2.0-3.9 | 1604 | 17.0% |
| White | 2799 | 29.6% | ≥4.0 | 551 | 5.8% |
| Black | 2757 | 29.2% | No data | 5438 | 57.6% |
| Asian | 568 | 6.0% | Change in Lactate |  |  |
| Hispanic | 2204 | 23.3% | Falling ≥0.2 | 2537 | 26.9% |
| Other* | 749 | 7.9% | Change <0.2 | 360 | 3.8% |
| Unknown/declined | 366 | 3.9% | Rising ≥0.2 | 1115 | 11.8% |
| Initial Temperature |  |  | No data | 5431 | 57.5% |
| 36.0-37.9C | 6431 | 68.1% | Blood Pressure (BP) |  |  |
| <36.0C | 720 | 7.6% | Systolic BP <90 | 2148 | 22.7% |
| >38.0C | 2175 | 23.0% | Diastolic BP <60 | 3126 | 33.1% |
| No data | 117 | 1.2% | Mean arterial pressure <65 | 1830 | 19.4% |
|  |  |  | Any of above | 3332 | 35.3% |

\* Includes American Indian, Alaskan/Hawaiian Native, Pacific Islander

Over two-thirds were initiated on the sepsis care pathway within one hour of arrival, and 95% within three hours. The median age was 71 years (interquartile range [IQR] 59-82), with nearly one third aged ≥80 years, and a slight majority were male. Racial and ethnic distribution were balanced: 29.6% White, 29.2% Black, and 23.3% Hispanic. Most patients were normothermic on arrival. Approximately, one-third presented with hypotension, and one-third had a normal WBC count (4,000-11,000/mL). Initial lactate was <2.0 mmol/L in a minority of patients but was <4.0 mmol/L in two-thirds. Among those who underwent repeat lactate testing, nearly two-thirds demonstrated a decrease of at least 0.2 mmol/L.

The 9,443 participants received 1,791 unique ED primary diagnoses. Table 2 summarizes the distribution of diagnostic categories based on our classification schema. Most participants were classified as having a non-infectious condition, followed in descending frequency by sepsis, infection, possible infection, and organ failure. Among ED primary diagnosis categorized as infection, the most common sources were pulmonary, genitourinary, skin and soft tissue, and gastrointestinal. Most remaining cases were febrile syndromes or unspecified infections (e.g., parasitic infection or abscess without specified location).

**Table 2:** Diagnostic Categories among ED Patients Placed on a Sepsis Care Pathway (n=9443) Table 2 describes categories of primary emergency department (ED) diagnoses in a cohort of 9,443 patients placed on a sepsis pathway in the ED of five hospitals in a major urban health system from 2022-2024, obtained from an electronic health record query. Categories were manually assigned by the prinicipal investigator using a schema modeled on that described by Linden and Melhammar.

|  | N (%) | % of Infection | % Severe*** | % Non-severe |
| --- | --- | --- | --- | --- |
| Sepsis | 1982 (21%) | -- | 25.6% | 0.0% |
| Serious Infection* | 365 (3.9%) | -- | 93.4% | 6.6% |
| Infection | 1874 (19.8%) | 100.0% | 6.8% | 86.9% |
| Pneumonia | 873 (9.2%) | 46.5% | 2.1% | 97.8% |
| GU Infection | 424 (4.5%) | 22.7% | 10.1% | 68.4% |
| GI Infection | 153 (1.6%) | 8.1% | 23.5% | 68.6% |
| SSTI** | 253 (2.7%) | 13.6% | 9.1% | 90.5% |
| ENT Infection | 15 (0.2%) | 0.8% | 13.3% | 80.0% |
| Febrile Syndrome | 130 (1.4%) | 7.1% | 0.8% | 98.5% |
| Unspecified/Other | 26 (0.3%) | 1.4% | 15.4% | 38.5% |
| Viral Infection | 256 (2.7%) | -- | 0.8% | 99.2% |
| Organ Failure | 1085 (11.5%) | -- | 97.8% | 0.5% |
| Non-infectious | 2667 (28.2%) | -- | 45.6% | 41.0% |
| Possible Infection | 1160 (12.3%) | -- | 51.2% | 32.8% |
| No Data | 54 (0.6%) | -- | -- | -- |

\*Infections associated with need for surgical intervention or high morbidity and mortality (e.g., CNS infection, intraabdominal infection, gangrene)
\*\*SSTI=Skin and soft tissue infection
\*\*\*Diagnoses generally associated with need for surgical intervention and/or hospitalization

### HLOS <72 hours, No ICU Stay, Survival to Discharge, and Sepsis Adjudication Outcome Frequency

The median HLOS was 194 hours (8.1 days; IQR 110-344), and increased with advancing age. Excluding deaths, 8.5% of participants had a short stay hospitalization (<72 hours). Most patients (65.9%) did not require ICU-level care, and 85% survived to hospital discharge. Additionally, more than one in four participants (28.0%) were adjudicated as not having sepsis.

### Multivariable Analysis of Patient Factors Linked to HLOS <72 hours, No ICU Stay, Survival to Discharge, and No Sepsis Adjudication Reference Standard Outcomes

Univariable associations for HLOS <72 hours, no ICU stay, survival to discharge, and adjudication as not having sepsis are presented in Supplement Tables S2A-S4B.

In the multivariable analysis (Table 3), several patient factors were independently associated with each reference-standard outcome.

**Table 3:** Multivariate Associations of Patient Factors with the Reference Standard Outcomes. Table 3 shows the adjusted odds ratios and 95% confidence intervals for associations between demographic and clinical factors and four reference outcomes in a cohort of 9,443 patients placed on a sepsis pathway in the emergency department of five hospitals in a major urban health system from 2022-2024. Data were obtained from an electronic health record query.

|  | HLOS <72 hours* | No ICU stay | Survival to discharge | No sepsis adjudication |
| --- | --- | --- | --- | --- |
|  | OR (95% CI) | OR (95% CI) | OR (95% CI) | OR (95% CI) |
| Age group |  |  |  |  |
| 70+ | Reference | Reference | Reference | Reference |
| 50-69 | 1.75 (1.45-2.12)<sup>††</sup> | 0.92 (0.83-1.03) | 1.68 (1.45-1.94)<sup>††</sup> | 1.25 (1.07-1.46) |
| 18-49 | 3.48 (2.81-4.29)<sup>††</sup> | 1.05 (0.90-1.22) | 3.15 (2.44-4.12)<sup>††</sup> | 1.01 (0.90-1.14) |
| Sex |  |  |  |  |
| Male | Reference | Reference | Reference | Reference |
| Female | 0.98 (0.83-1.15) | 0.95 (0.86-1.05) | 1.05 (0.92-1.19) | 1.10 (0.99-1.22) |
| Race |  |  |  |  |
| Black or other | Reference | Reference | Reference | Reference |
| Not Black or other | 1.56 (1.30-1.88)<sup>††</sup> | 1.16 (1.04-1.28)<sup>†</sup> | 1.14 (1.0-1.30) | 1.02 (0.91-1.14) |
| Temperature |  |  |  |  |
| <36C | Reference | Reference | Reference | Reference |
| ≥36C | 1.61 (1.10-2.47)<sup>†</sup> | 1.97 (1.65-2.36)<sup>††</sup> | 2.28 (1.87-2.76)<sup>††</sup> | 1.01 (0.83-1.22) |
| Hypotension |  |  |  |  |
| Any | Reference | Reference | Reference | Reference |
| None | 1.22 (1.01-1.47)<sup>†</sup> | 1.47 (1.33-1.63)<sup>††</sup> | 1.48 (1.30-1.68)<sup>††</sup> | 1.07 (0.96-1.20) |
| WBC count |  |  |  |  |
| <4.0K or ≥15.0K | Reference | Reference | Reference | Reference |
| 4.0K-14.9K | 1.24 (1.05-1.47)<sup>†</sup> | 1.29 (1.17-1.42)<sup>††</sup> | 1.55 (1.36-1.76)<sup>††</sup> | 1.70 (1.53-1.89)<sup>††</sup> |
| Lactate |  |  |  |  |
| High | Reference | Reference | Reference | Reference |
| Low** | 0.98 (0.82-1.17) | 1.25 (1.13-1.39)<sup>††</sup> | 2.09 (1.79-2.44)<sup>††</sup> | 1.25 (1.12-1.40)<sup>††</sup> |
| ED infection or sepsis diagnosis |  |  |  |  |
| Sepsis | Reference | Reference | Reference | Reference |
| Pneumonia | 1.31 (0.92-1.84) | 1.41 (1.16-1.72)<sup>††</sup> | 0.95 (0.74-1.23) | 1.73 (1.29-4.55)<sup>††</sup> |
| Any other infection | 1.47 (1.11-1.96)<sup>†</sup> | 2.72 (2.30-3.23)<sup>††</sup> | 2.20 (1.73-2.81)<sup>††</sup> | 3.61 (2.88-4.55)<sup>††</sup> |
| No infection/no sepsis | 1.85 (1.45-2.38)<sup>††</sup> | 1.44 (1.26-1.63)<sup>††</sup> | 1.05 (0.89-1.24) | 8.82 (7.23-10.84)<sup>††</sup> |
| ED diagnosis severity |  |  |  |  |
| Severe | Reference | Reference | Reference | Reference |
| All others | 1.42 (1.19-1.71)<sup>††</sup> | 2.74 (2.46-3.05)<sup>††</sup> | 1.82 (1.58-2.09)<sup>††</sup> | 1.01 (0.90-1.13) |

\*Excluding patients who were admitted and expired within 72 hours
\*\*Repeat lactate <2.0 mmol/L, or if no repeat measured, initial lactate <2.0 mmol/L
<sup>†</sup>p<0.05 <sup>††</sup>p<0.01

For HLOS <72 hours, associated factors included age ≤49 years, race other than Black or “other”, initial temperature ≥36° C, absence of hypotension, WBC count 4,000/uL to 15,000/uL, an ED diagnosis of infection other than pneumonia or non-infectious/non-sepsis diagnosis, and absence of a severe ED diagnosis.

For no ICU stay, associated factors included Black or other race, initial temperature ≥36° C, absence of hypotension, WBC count 4,000/uL to 15,000/uL, repeat lactate testing or a single lactate ≥2.0mmol/L, an ED diagnosis of infection other than pneumonia, and absence of severe ED diagnosis.

For survival to discharge, associated factors were age ≤49 years, initial temperature ≥36° C, absence of hypotension, WBC count 4,000/uL to 15,000/uL, an ED diagnosis of infection other than sepsis or pneumonia, and absence of severe ED diagnosis.

For not being adjudicated as having sepsis, associated factors were age ≤49 years, WBC count 4,000/uL to 15,000/uL, repeat lactate ≥2.0mmol/L or a single measured lactate ≥2.0mmol/L, and an ED diagnosis of infection other than pneumonia or non-sepsis/non-infection diagnoses.

## DISCUSSION

Among patients initiated on a sepsis care pathway, our goal was to identify patient factors associated with more favorable clinical and operational outcomes – specifically HLOS <72 hours, no ICU stay, survival to discharge, and adjudication of not having sepsis - that might help identify candidates for sepsis care de-escalation. The factors associated with these outcomes varied depending on the reference standard used, although several factors were common across multiple outcomes. Younger age was associated with short hospital stay, survival, and adjudication as not having sepsis. Initial temperature ≥36° C, absence of hypotension, and absence of severe ED diagnosis were each associated with short stay, survival, and avoiding ICU stay. Normal lactate levels were linked to avoiding ICU care, survival, and not being adjudicated as having sepsis. An ED diagnosis of infection (excluding pneumonia) or a non-infection/non-sepsis diagnosis was associated with both no ICU stay and survival to discharge. A severe ED diagnosis was associated with HLOS <72 hours, no ICU stay, and survival. Only one factor - WBC count 4,000/uL to 15,000/uL - was associated with all four reference outcomes.

These results suggest that several readily identifiable patient factors and ED diagnostic impressions may help flag patients on a sepsis pathway who are at lower risk of adverse outcomes. However, additional research is needed before these factors can be reliably used to guide sepsis care de-escalation. We see several necessary next steps for advancing this work. First, additional objective patient factors and subjective clinician assessments must be evaluated prospectively, to capture information not routinely available through retrospective EHR review in structured form. Second, it will be essential to determine which combinations of factors most efficiently identify low-risk patients, the proportion of patients on a sepsis pathway who meet these combined criteria, and the absolute risk of each reference outcome among those identified. It will also be important to determine whether a combination of factors can consistently identify low-risk patients across all reference outcomes. Third, future studies should evaluate the test performance of these factors – including sensitivity, specificity, predictive value, receiver operator characteristic (ROC) area under the curve (AUC), and likelihood ratios - and validate their performance in prospective cohorts. Finally, further work is needed to determine whether these factors can enhance interpretation and clinical application of emerging diagnostic assays (e.g., Intellisep® (Cytovale®, San Francisco, CA), monocyte distribution width (MDW, Beckman Coulter, Brea, CA), Triverity™ (Inflammatix™, Sunnyvale, CA) and to examine longer-term patient outcomes.

The identification of patients whose sepsis care might be safely de-escalated during their ED visit has been understudied, leaving few published investigations for comparison with our findings. Mulders et. al. conducted a prospective study at a single academic hospital in the Netherlands from 2016 to 2019 (annual patient volume 34,000), enrolling adult patients with suspected infection and at least two SIRS criteria. ^29^ In contrast to our study, patients admitted to the ICU were excluded, preventing ICU stay from being evaluated as an outcome. The investigators’ primary objective was to identify clinical factors associated with “safe early discharge”, defined as discharge from the hospital within 24 hours and no disease-related readmission or death within seven days. Factors associated with “safe early discharge” included younger age, absence of comorbidities, living independently, lower triage acuity, not arriving by ambulance, not being referred by a general practitioner, normal clinical impression and risk scores (i.e., qSOFA, PIRO, MEDS, NEWS, and SIRS), normal vital signs, and absence of kidney or respiratory failure. In contrast to our investigation, Mulders et al. used a single reference standard (“safe early discharge”), and many of the factors they identified reflect information obtained during the course of hospitalization rather than data available during the ED visit.

It is worth noting several ways in which our findings align with prior studies. Our cohort included a similar proportion of older adults - approximately half of participants were aged ≥70 years – consistent with earlier sepsis investigations. ^30,31^ Outcome frequencies in our study were also comparable to those reported in a large national analysis by Kuye et al:.— short-stay admission < 72 hours (8.5% vs. 10.2%), ICU admission (34.1% vs. 29%), and in-hospital mortality (15.0% vs. 13.1%).^32^ Similar to prior studies, we also found that the absence of hypothermia was associated with lower mortality and a lower likelihood of being adjudicated as having sepsis.^42,43^ Our finding that Black race was associated with less favorable outcomes is consistent with previous studies and warrants further investigation. A 2023 meta-analysis reported that Black patients often face higher sepsis-related mortality rates (approx. 20-30% higher in some regions) compared to White counterparts, even when adjusting for comorbidities.^33^ These disparities likely reflect a complex interplay of structural, socioeconomic, and healthcare-system factors rather than biologic differences, ^34^ and highlight the importance of equity-focused research in sepsis care.

### Limitations

Our study has several limitations. First, by restricting our analysis to patients already placed on a sepsis care pathway, we likely selected a cohort that was more overtly ill. This may limit the generalizability of our findings to the broader population of ED patients being evaluated for possible sepsis. Nonetheless, the characteristics and outcomes of our cohort are consistent with those reported in prior work (e.g., Singer et al., 2016).^35^

Second, the retrospective nature of our study prevented us from assessing factors not captured in the EHR. We also chose not to include certain available variables - such as heart rate, respiratory rate, and oxygen saturation - because these tend to be variable over the course of the ED visit, are less specific indicators of infection, and in the case of oxygen saturation, may be confounded by inaccurate documentation of oxygen delivery. Additionally, our decision to categorize variables into binary or categorical groups, mirroring how ED clinicians typically synthesize information, may have obscured more nuanced associations, including threshold or non-linear effects.

Third, our ED diagnosis categories were assigned based on the principal investigator’s clinical judgment and should be interpreted as exploratory. We were unable to identify a validated classification system based solely on ED diagnosis. Because only the primary diagnosis was extracted, some patients may have had both infectious and non-infectious conditions that were not captured. Given that our goal was to identify factors that could support de-escalation of sepsis care, the presence of a substantial number of sepsis mimics – recognized during the course of ED work-up – could limit the applicability of our findings to patients for whom sepsis remains a concern at the time of ED disposition.

Fourth, alternative reference standard outcomes are possible and could better reflect the safety of de-escalating sepsis care. As our results illustrate, different outcomes definitions likely would yield further variation in the patient factors that identify candidates for de-escalation of sepsis care.

## CONCLUSION

Several patient factors were associated with favorable clinical outcomes - defined as hospital length of stay under 72 hours, no ICU admission, survival to discharge, and adjudication as not having sepsis. In combination, these factors may help identify patients who could be appropriate candidates for de-escalation of sepsis care. Prospective studies are needed to determine which combination of predictive factors most effectively guide safe de-escalation strategies for low-risk patients.

## Data Availability

All data produced in the present study are available upon reasonable request to the authors.

## Acknowledgements

Cindy Mei, MS assisted with performing the electronic health record data query.

## Author contributions

Study concept and design (BW, SES, ML, NIS, RCM); Acquisition of data (BW, CY); Analysis and interpretation of the data (BW, CY, MM, RCM); Drafting of the manuscript (BW, ASG); Critical revision of the manuscript for important intellectual content (SES, ML, NIS, MM, RCM); Statistical expertise (CY, MM); Acquisition of funding (N/A)

