## Supplemental Figure and Tables for "Low-risk Features of Emergency Department Patients on a Sepsis Pathway: A Retrospective Cohort Study"

### SUPPLEMENT

Figure S1: STROBE Statement—Checklist of items that should be included in reports of ***cohort studies***

| | Item No | Recommendation | Page No |
|---|---|---|---|
| **Title and abstract** | 1 | (*a*) Indicate the study's design with a commonly used term in the title or the abstract<br><br>(*b*) Provide in the abstract an informative and balanced summary of what was done and what was found | 1 |
| **Introduction** | | | |
| Background/rationale | 2 | Explain the scientific background and rationale for the investigation being reported | 5 |
| Objectives | 3 | State specific objectives, including any prespecified hypotheses | 7 |
| **Methods** | | | |
| Study design | 4 | Present key elements of study design early in the paper | 8 |
| Setting | 5 | Describe the setting, locations, and relevant dates, including periods of recruitment, exposure, follow-up, and data collection | 8 |
| Participants | 6 | (*a*) Give the eligibility criteria, and the sources and methods of selection of participants. Describe methods of follow-up<br><br>(*b*) For matched studies, give matching criteria and number of exposed and unexposed | 8 |
| Variables | 7 | Clearly define all outcomes, exposures, predictors, potential confounders, and effect modifiers. Give diagnostic criteria, if applicable | 9 |
| Data sources/ measurement | 8* | For each variable of interest, give sources of data and details of methods of assessment (measurement). Describe comparability of assessment methods if there is more than one group | 9 |
| Bias | 9 | Describe any efforts to address potential sources of bias | n/a |
| Study size | 10 | Explain how the study size was arrived at | n/a |
| Quantitative variables | 11 | Explain how quantitative variables were handled in the analyses. If applicable, describe which groupings were chosen and why | 10 |
| Statistical methods | 12 | (*a*) Describe all statistical methods, including those used to control for confounding<br>(*b*) Describe any methods used to examine subgroups and interactions<br>(*c*) Explain how missing data were addressed<br>(*d*) If applicable, explain how loss to follow-up was addressed<br>(*e*) Describe any sensitivity analyses | 10 |
| **Results** | | | |
| Participants | 13* | (a) Report numbers of individuals at each stage of study—e.g., numbers potentially eligible, examined for eligibility, confirmed eligible, included in the study, completing follow-up, and analyzed | |

| | | | |
|---|---|---|---|
| | | (b) Give reasons for non-participation at each stage<br>(c) Consider use of a flow diagram | 11 |
| Descriptive data | 14* | (a) Give characteristics of study participants (e.g., demographic, clinical, social) and information on exposures and potential confounders<br>(b) Indicate number of participants with missing data for each variable of interest<br>(c) Summarize follow-up time (e.g., average and total amount) | 12 |
| Outcome data | 15* | Report numbers of outcome events or summary measures over time | 13 |
| Main results | 16 | (*a*) Give unadjusted estimates and, if applicable, confounder-adjusted estimates and their precision (e.g., 95% confidence interval). Make clear which confounders were adjusted for and why they were included<br>(*b*) Report category boundaries when continuous variables were categorized<br>(*c*) If relevant, consider translating estimates of relative risk into absolute risk for a meaningful time period | 13-15 |
| Other analyses | 17 | Report other analyses done—e.g., analyses of subgroups and interactions, and sensitivity analyses | n/a |
| **Discussion** | | | |
| Key results | 18 | Summarize key results with reference to study objectives | 16 |
| Limitations | 19 | Discuss limitations of the study, taking into account sources of potential bias or imprecision. Discuss both direction and magnitude of any potential bias | 19 |
| Interpretation | 20 | Give a cautious overall interpretation of results considering objectives, limitations, multiplicity of analyses, results from similar studies, and other relevant evidence | 20 |
| Generalizability | 21 | Discuss the generalizability (external validity) of the study results | 18 |
| **Other information** | | | |
| Funding | 22 | Give the source of funding and the role of the funders for the present study and, if applicable, for the original study on which the present article is based | n/a |

*Give information separately for exposed and unexposed groups.

**Note:** An Explanation and Elaboration article discusses each checklist item and gives methodological background and published examples of transparent reporting. The STROBE checklist is best used in conjunction with this article (freely available on the Web sites of PLoS Medicine at http://www.plosmedicine.org/, Annals of Internal Medicine at http://www.annals.org/, and Epidemiology at http://www.epidem.com/). Information on the STROBE Initiative is available at http://www.strobe-statement.org.

Table S1: Frequent and Representative Examples of ED Diagnosis by Assigned Categories

| Category | Most frequent and representative diagnoses (frequency) |
|---|---|
| Infection | |
| Pneumonia | Pneumonia due to infectious organism, unspecified laterality (183); Pneumonia (173); Multifocal pneumonia (122); Pneumonia of right lower lobe due to infectious organism (69); Bacterial pneumonia (57) |
| GU infection | Pyelonephritis (108); Urinary tract infection without hematuria, site unspecified (91); Urinary tract infection, site not specified (57); UTI (urinary tract infection) (36); Urinary tract infection with hematuria, site unspecified (32) |
| GI infection | Colitis (45); Gastroenteritis (20); Cholangitis (17); Stercoral colitis (12); Enterocolitis (8) |
| SSTI | Cellulitis (33); Cellulitis of left lower extremity (23); Cellulitis of right lower extremity (20); Cellulitis and abscess of leg (19); Cellulitis of lower extremity, unspecified laterality (12) |
| Fever | Fever, unspecified fever cause (89); Fever (31); Fever of unknown origin (17); Fever and chills (9); Fever, unknown origin (5) |
| ENT infection | Strep pharyngitis (7); peritonsillar abscess (3); strep throat (3); acute pharyngitis, unspecified etiology (3); dental infection (2) |
| Other/unspecified Infection | Malaria (4); wound infection (4); infection (3); post-op infection (2); MRSA infection (2) |
| Severe infection | Neutropenic fever (48); acute cholecystitis (44); bacteremia (17); cholecystitis (14); diverticulitis of colon (13); bowel perforation (10); osteomyelitis (10); liver abscess (6); Acute bacterial endocarditis (6); gangrene (6); meningitis (4) |
| Non-infectious | Cardiac arrest (144); Seizure (58); Abdominal pain, unspecified abdominal location (55); Syncope and collapse (51); Hypernatremia (50); Obstructive chronic bronchitis with exacerbation (49); SBO (small bowel obstruction (48); Pleural effusion (43) |
| Possible infection | Hypotension, unspecified hypotension type (176); Shortness of breath (162); Hypotension (97); Failure to thrive in adult (62); Respiratory distress (43); Weakness (38); Diarrhea, unspecified type (19) |
| Organ Failure | Altered mental status, unspecified altered mental status type (226); Acute respiratory failure with hypoxia (193); Hypoxia (156); AKI (acute kidney injury) (141); Acute hypoxic respiratory failure (53); Acute respiratory failure with hypoxia and hypercapnia (49); Shock (45) |
| Sepsis | Sepsis, due to unspecified organism, unspecified whether acute organ dysfunction present (650); Sepsis (471); Septic shock (363); Severe sepsis (330); Sepsis without acute organ dysfunction, due to unspecified organism (84) |
| Viral infection | COVID-19 (169); Influenza A (55); Influenza (31); Acute viral syndrome (17); Respiratory syncytial virus (RSV) (8); Upper respiratory tract infection, unspecified type (5) |

| | |
|---|---|
| Non-severe | Pneumonia (478); Shortness of breath (162); COVID-19 (106); Urinary tract infection (91); Fever, unspecified (89); Seizure (58); Abdominal pain, site unspecified (55) |
| Severe | Septic shock (364); altered mental status, unspecified type (226); Acute respiratory failure with hypoxia (193); Hypotension, unspecified type (176); Hypoxia (156); Cardiac arrest (144); Acute kidney injury (141) |
| Other | Sepsis due to unspecified organism, unspecified whether acute organ dysfunction present (650); Sepsis (471); Severe sepsis (330); Pyelonephritis (108); Syncope and collapse (51); Hyponatremia (38); Elevated troponin (17) |

Table S1 shows examples of frequent and representative emergency department diagnoses within categories modeled on the schema of Linden and Melhammar. Categories were assigned manually by the principal investigator using data obtained in an electronic health record query on 9,443 patients placed on a sepsis pathway.

Table S2-A: Univariable Associations of Demographic Factors with Outcomes of Patients Placed on Sepsis Pathway

| | Hospital LOS* | | | No ICU stay | |
|---|---|---|---|---|---|
| | Median (IQR) | <72 hours | Odds ratio (95% CI) | % | OR** |
| All patients | 194 (110–344) | 8.5% | -- | 65.9% | |
| Age | | | | | |
| 18-29 | 119 (75–210) | 23.7% | reference | 68.9% | reference |
| 30-39 | 142 (81–279) | 20.4% | 0.82 (0.56-1.22) | 72.0% | 1.16 (0.82-1.65) |
| 40-49 | 165 (98–308) | 14.4% | 0.54 (0.37-0.80)†† | 69.9% | 1.05 (0.75-1.45) |
| 50-59 | 187 (109–344) | 9.1% | 0.32 (0.22-0.47)†† | 64.0% | 0.80 (0.59-1.09) |
| 60-69 | 197 (108–363) | 9.5% | 0.34 (0.24-0.48)†† | 65.1% | 0.84 (0.62-1.13) |
| 70-79 | 207 (120–368) | 6.7% | 0.23 (0.16-0.33)†† | 63.4% | 0.78 (0.58-1.05) |
| 80-89 | 209 (126–350) | 4.9% | 0.16 (0.11-0.24)†† | 65.0% | 0.84 (0.62-1.12) |
| 90-99 | 198 (116–310) | 4.3% | 0.15 (0.09-0.23)†† | 71.5% | 1.13 (0.82-1.55) |
| Age group | | | | | |
| 18-49 | 148 (85-284) | 18.1% | reference | 70.4% | reference |
| 50-69 | 193 (108-357) | 9.3% | 0.47 (0.39-0.56)†† | 64.7% | 0.77 (0.67-0.88)†† |
| 70+ | 205 (123-348) | 5.6% | 0.27 (0.22-0.33)†† | 65.4% | 0.79 (0.70-0.91)†† |
| Sex | | | | | |
| Male | 195 (111–349) | 8.9% | reference | 66.5% | reference |
| Female | 193 (110–337) | 8.3% | 0.93 (0.80-1.08) | 65.2% | 0.94 (0.86-1.03) |
| Race | | | | | |
| Asian | 163 (98–293) | 11.5% | reference | 66.4% | reference |
| White | 191 (109–335) | 9.0% | 0.77 (0.57-1.04) | 64.2% | 0.91 (0.75-1.10) |
| Hispanic | 185 (105–326) | 9.6% | 0.82 (0.61-1.12) | 70.6% | 1.22 (1.00-1.48) |
| Black/African American | 206 (121–366) | 6.9% | 0.57 (0.43-0.78)†† | 63.7% | 0.89 (0.73-1.08) |
| Other | 216 (117–367) | 7.4% | 0.62 (0.42-0.91)† | 62.1% | 0.83 (0.66-1.04) |
| Unknown /declined | 204 (105–367) | 9.5% | 0.81 (0.52-1.26) | 73.8% | 1.42 (1.07-1.91)† |

*ED LOS plus Inpatient LOS for patients who did not expire; excludes patients who died within 72 hours

**For each category, odds ratio and p-value relative to lowest risk group

† $p < 0.05$ †† $p < 0.01$

Table S2-A shows univariable odds ratios and 95% confidence intervals for associations between demographic factors and two reference outcomes (hospital length of stay <72 hours and no ICU stay) in a cohort of 9,443 patients placed on a sepsis pathway in the emergency department of five hospitals in a major urban health system from 2022-2024. Data were obtained from an electronic health record query.

Table S2-B: Univariable Associations of Demographic Factors with Outcomes of Patients Placed on Sepsis Pathway

| | Survival to discharge | | Not sepsis adjudication | |
|---|---|---|---|---|
| | % | OR** | % | OR** |
| All patients | 84.5% | | 28.0% | -- |
| Age | | | | |
| 18-29 | 98.2% | reference | 29.8% | reference |
| 30-39 | 95.1% | 0.35 (0.10-0.93) | 34.5% | 1.23 (0.88-1.75) |
| 40-49 | 91.5% | 0.20 (0.06-0.49)†† | 31.1% | 1.06 (0.76-1.49) |
| 50-59 | 87.9% | 0.13 (0.04-0.32)†† | 27.6% | 0.90 (0.66-1.23) |
| 60-69 | 87.1% | 0.12 (0.04-0.29)†† | 28.4% | 0.93 (0.69-1.27) |
| 70-79 | 83.2% | 0.09 (0.03-0.21)†† | 25.7% | 0.81 (0.61-1.11) |
| 80-89 | 79.9% | 0.07 (0.02-0.17)†† | 28.2% | 0.93(0.69-1.27) |
| 90-99 | 74.3% | 0.05 (0.02-0.12)†† | 26.7% | 0.86 (0.63-1.19) |
| Age group | | | | |
| 18-49 | 93.9% | reference | 32.0% | reference |
| 50-69 | 87.4% | 0.45 (0.35-0.57)†† | 28.1% | 0.83 (0.72-0.95)† |
| 70+ | 80.4% | 0.26 (0.21-0.33)†† | 26.8% | 0.78 (0.68-0.88)†† |
| Sex | | | | |
| Male | 85.0% | reference | 27.0% | reference |
| Female | 83.9% | 0.92 (0.82-1.02) | 29.1% | 1.11 (1.01-1.22)† |
| Race | | | | |
| Asian | 84.7% | reference | 28.0% | reference |
| White | 83.4% | 0.91 (0.70-1.16) | 26.8% | 0.94 (0.78-1.15) |
| Hispanic | 86.8% | 1.19 (0.91-1.54) | 27.9% | 1.00 (0.81-1.23) |
| Black/African American | 84.1% | 0.96 (0.74-1.23) | 28.1% | 1.00 (0.82-1.23) |
| Other | 82.4% | 0.85 (0.63-1.13) | 28.7% | 1.03 (0.81-1.32) |
| Unknown/declined | 85.5% | 1.07 (0.74-1.55) | 34.2% | 1.33 (1.00-1.79) |

*ED LOS plus Inpatient LOS for patients who did not expire; excludes patients who died within 72 hours

**For each category, odds ratio and p-value relative to lowest risk group

† $p < 0.05$ †† $p < 0.01$

Table S2-B shows univariable odds ratios and 95% confidence intervals for associations between demographic factors and two reference outcomes (survival to discharge and not being adjudicated as having sepsis by the healthcare system's institutional sepsis committee) in a cohort of 9,443 patients placed on a sepsis pathway in the emergency department of five hospitals in a major urban health system from 2022-2024. Data were obtained from an electronic health record query.

Table S3-A: Univariable Associations of Clinical Factors with Outcomes of Patients Placed on Sepsis Pathway

| | Hospital LOS* | | | No ICU stay | |
|---|---|---|---|---|---|
| | Median (IQR) | <72 hours | Odds ratio (95% CI) | % | OR** |
| Fever | | | | | |
| Initial Temp 36-37.9C | 196 (112–343) | 8.9% | reference | 66.2% | reference |
| Initial Temp >38C | 184 (110–323) | 8.7% | 0.97 (0.81-1.15) | 72.2% | 1.33 (1.19-1.48)†† |
| Initial Temp<36C | 209 (99–399) | 5.3% | 0.57 (0.39-0.81)† | 48.2% | 0.47 (0.41-0.55)†† |
| No data | 223 (97–435) | 8.2% | 0.91 (0.40-1.77) | 37.6% | 0.31 (0.21-0.45)†† |
| Hypotension | | | | | |
| No hypotension | 191 (109–333) | 9.4% | reference | 69.7% | reference |
| Initial SBP <90 | 195 (106–350) | 7.7% | 0.86 (0.71-1.03) | 57.1% | 0.61 (0.55-0.68)†† |
| Initial DBP <60 | 200 (114–360) | 7.1% | 0.74 (0.63-0.88)†† | 58.7% | 0.62 (0.57-0.68)†† |
| Initial MAP < 65 | 198 (108–366) | 7.5% | 0.84 (0.69-1.02)† | 55.1% | 0.57 (0.51-0.63)†† |
| Initial WBC count | | | | | |
| 4.0 - 10.9K | 184 (105–325) | 9.7% | reference | 68.6% | reference |
| <4.0 K | 187 (109–343) | 7.2% | 0.72 (0.51-0.99) | 63.1% | 0.79 (0.66-0.94)† |
| 11.0 - 14.9K | 193 (110–337) | 9.0% | 0.93 (0.76-1.13) | 67.5% | 0.95 (0.84-1.07) |
| 15.0 -19.9K | 200 (114–355) | 8.0% | 0.82 (0.65-1.01) | 64.2% | 0.82 (0.73-0.93)†† |
| ≥20.0 K | 205 (117–367) | 7.3% | 0.74 (0.59-0.91)† | 62.5% | 0.76 (0.68-0.86)†† |
| 4.0-14.9K | 187 (106-331) | 9.4% | reference | 68.1% | reference |
| <4.0K or ≥15.0K | 201 (115-361) | 7.6% | 0.79 (0.68-0.92)†† | 63.3% | 0.81 (0.74-0.88)†† |
| Initial Lactate | | | | | |
| <2.0 | 198 (110–351) | 9.2% | reference | 69.3% | reference |
| 2.0-3.9 | 194 (116–337) | 8.5% | 0.92 (0.75-1.12) | 71.2% | 1.09 (0.97-1.23) |
| ≥4.0 | 191 (99–348) | 8.6% | 0.93 (0.74-1.16) | 54.1% | 0.52 (0.46-0.59)†† |
| No data | 199 (126–378) | 5.7% | 0.60 (0.25-1.23) | 66.1% | 0.86 (0.59-1.28) |
| Repeat Lactic acid | | | | | |
| <2.0 | 207 (126–352) | 8.1% | reference | 69.1% | reference |
| 2.0-3.99 | 206 (123–353) | 7.7% | 0.94 (0.73-1.21) | 71.0% | 1.09 (0.94-1.27) |
| ≥4.0 | 198 (83–373) | 6.6% | 0.80 (0.52-1.18) | 43.0% | 0.34 (0.28-0.41)†† |
| No data | 186 (104–331) | 9.2% | 1.14 (0.94-1.39) | 65.6% | 0.85 (0.76-0.95)† |
| Change in lactate | | | | | |
| Change < 0.2 | 209 (118–372) | 6.2% | reference | 66.7% | reference |
| Rising ≥0.2 | 194 (112–351) | 8.2% | 1.36 (0.84-2.28) | 66.4% | 0.99 (0.77-1.27) |
| Falling ≥0.2 | 208 (124–359) | 7.8% | 1.29 (0.83-2.12) | 66.2% | 0.98 (0.77-1.24) |
| Missing data | 186 (104–331) | 9.2% | 1.54 (1.01-2.50) | 65.6% | 0.95 (0.76-1.19) |

| | | | | | |
|---|---|---|---|---|---|
| Repeat lactate <2 or if no repeat, initial <2 | 207 (126–352) | 8.1% | reference | 69.1% | reference |
| All other lactate | 191 (107-341) | 8.6% | 1.06 (0.88-1.28) | 64.4% | 0.81 (0.72-0.90)†† |

*ED LOS plus Inpatient LOS for patients who did not expire; excludes patients who died within 72 hours
**For each category, odds ratio and p-value relative to lowest risk group
† $p < 0.05$ †† $p < 0.01$

Table S3-A shows univariable odds ratios and 95% confidence intervals for associations between clinical factors and two reference outcomes (hospital length of stay <72 hours and no ICU stay) in a cohort of 9,443 patients placed on a sepsis pathway in the emergency department of five hospitals in a major urban health system from 2022-2024. Data were obtained from an electronic health record query.

Table S3-B: Univariable Associations of Clinical Factors with Outcomes of Patients Placed on Sepsis Pathway

| | Survival to discharge | | Not sepsis adjudication | |
|---|---|---|---|---|
| | % | OR** | % | OR** |
| Fever | | | | |
| Initial Temp 36-37.9C | 84.6% | reference | 30.1% | reference |
| Initial Temp >38C | 90.4% | 1.71 (1.46-2.00)†† | 18.8% | 0.53 (0.47-0.60)†† |
| Initial Temp<36C | 68.3% | 0.39 (0.33-0.47)†† | 34.3% | 1.20 (1.03-1.43)† |
| No data | 65.0% | 0.34 (0.23-0.50)†† | 41.0% | - |
| Hypotension | | | | |
| No hypotension | 86.6% | reference | 28.4% | reference |
| Initial SBP <90 | 80.2% | 0.67 (0.59-0.76)†† | 30.0% | 1.14 (1.02-1.27)† |
| Initial DBP <60 | 80.5% | 0.65 (0.58-0.72)†† | 27.0% | 0.93 (0.85-1.02) |
| Initial MAP < 65 | 79.3% | 0.64 (0.57-0.71)†† | 29.3% | 1.09 (0.97-1.22) |
| Initial WBC count | | | | |
| 4.0 - 10.9K | 86.8% | reference | 36.2% | reference |
| <4.0 K | 77.4% | 0.52 (0.42-0.64)†† | 23.9% | 0.55 (0.46-0.67)†† |
| 11.0 - 14.9K | 86.9% | 1.01 (0.85-1.19) | 30.4% | 0.77 (0.68-0.86)†† |
| 15.0 -19.9K | 84.1% | 0.80 (0.68-0.95)† | 23.1% | 0.53 (0.46-0.61)†† |
| ≥20.0 K | 81.2% | 0.65 (0.56-076)†† | 18.2% | 0.39 (0.34-0.45)†† |
| 4.0-14.9K | 86.8% | reference | | reference |
| <4.0K or ≥15.0K | 81.7% | 0.68 (0.61-0.76)†† | | 0.54 (0.49-0.60)†† |
| Initial Lactate | | | | |
| <2.0 | 89.7% | reference | 33.1% | reference |
| 2.0-3.9 | 87.8% | 0.83 (0.69-0.99)† | 24.5% | 0.66 (0.58-0.74)†† |
| ≥4.0 | 75.2% | 0.35 (0.29-0.42)†† | 31.2% | 0.92 (0.81-1.04) |
| No data | 82.7% | 0.55 (0.34-0.91)† | 25.2% | - |
| Repeat Lactic acid | | | | |
| <2.0 | 90.3% | reference | 29.3% | reference |
| 2.0-3.99 | 85.5% | 0.64 (0.52-0.78)†† | 25.0% | 0.81 (0.69-0.93)†† |
| ≥4.0 | 60.4% | 0.16 (0.13-0.21)†† | 26.3% | 0.86 (0.69-1.06) |
| No data | 84.6% | 0.59 (0.50-0.70)†† | 28.5% | - |
| Change in lactate | | | | |
| Change < 0.2 | 80.0% | reference | 25.6% | reference |
| Rising ≥0.2 | 80.4% | 1.02 (0.76-1.37) | 28.0% | 1.14 (0.87-1.49) |
| Falling ≥0.2 | 86.6% | 1.62 (1.22-2.14)†† | 27.1% | 1.09 (0.85-1.41) |
| Missing data | 84.6% | 1.37 (1.04-1.79)† | 28.5% | - |
| Repeat lactate <2 or if no repeat, initial <2 | 90.3% | reference | 29.3% | reference |
| All other lactate | 81.9% | 0.49 (0.41-0.57)†† | 26.5% | 0.87 (0.78-0.98)† |

*ED LOS plus Inpatient LOS for patients who did not expire; excludes patients who died within 72 hours

**For each category, odds ratio and p-value relative to lowest risk group

† $p < 0.05$ †† $p < 0.01$

Table S3-B shows univariable odds ratios and 95% confidence intervals for associations between clincal factors and two reference outcomes (survival to discharge and not being adjudicated as having sepsis by the healthcare system's institutional sepsis committee) in a cohort of 9,443 patients placed on a sepsis pathway in the emergency department of five hospitals in a major urban health system from 2022-2024. Data were obtained from an electronic health record query.

Table S4-A: Univariable Associations of Diagnoses with Outcomes of Patients Placed on Sepsis Pathway

| | Hospital LOS* | | | No ICU Stay | |
|---|---|---|---|---|---|
| | Median (IQR) | <72 hours | OR | % | OR |
| All patients | 193.9 (110.0–343.9) | 8.5% | -- | 65.9% | -- |
| Sepsis | 209.6 (123.0–375.6) | 5.4% | 0.57 (0.44-0.74)†† | 60.3% | 0.40 (0.34-0.46)†† |
| Serious Infection** | 212.7 (125.5–385.3) | 8.4% | 0.92 (0.60-1.36) | 73.4% | 0.72 (0.56-0.93)† |
| Infection | 182.4 (110.2–315.1) | 9.1% | reference | 79.3% | reference |
| Pneumonia | 201.8 (115.6–363.8) | 8.4% | reference*** | 74.1% | reference*** |
| GU infection | 167.8 (109.6–270.8) | 9.3% | 1.12 (0.73-1.68) | 82.5% | 1.65 (1.24-2.23)†† |
| GI infection | 153.5 (98.1–270.5) | 14.3% | 1.82 (1.06-3.02)† | 82.4% | 1.63 (1.06-2.58)† |
| SSTI | 188.6 (126.7–317.8) | 8.5% | 1.02 (0.60-1.67) | 85.4% | 2.04 (1.41-3.02)†† |
| ENT infection | 134.4 (86.1–233.2) | 21.4% | 2.98 (0.66-9.80) | 93.3% | 4.89 (0.98-88.87) |
| Fever | 159.3 (106.3–251.6) | 7.9% | 0.93 (0.44-1.78) | 86.2% | 2.17 (1.32-3.77)†† |
| Unspecified/other | 274.8 (194.6–471.5) | 3.8% | 0.44 (0.02-2.11) | 84.6% | 1.92 (0.73-6.62) |
| Viral Infection | 169.3 (96.7–276.8) | 10.6% | 1.18 (0.74-1.81) | 76.2% | 0.83 (0.61-1.14) |
| Organ Failure | 226.9 (119.5–401.3) | 6.9% | 0.74 (0.55-0.98)† | 50.5% | 0.27 (0.23-0.31)†† |
| Non-infectious | 182.1 (101.0–318.9) | 10.7% | 1.20 (0.98-1.48) | 64.6% | 0.48 (0.41-0.54)†† |
| Possible infection | 189.8 (105.5–338.9) | 9.0% | 0.99 (0.76-1.29) | 65.6% | 0.50 (0.42-0.59)†† |
| Non-severe | 181.0 (105.8–315.0) | 10.2% | reference | 77.8% | Reference |
| Severe dx**** | 209.2 (114.4–372.2) | 7.2% | 0.69 (0.58-0.82)†† | 52.2% | 0.31 (0.28-0.35)†† |
| Other | 191.6 (112.2–342.6) | 8.2% | 0.79 (0.65-0.95)† | 71.2% | 0.71 (0.62-0.80)†† |
| Boarding (ED LOS >24 hours) | 209.9 (135.1–345.1) | 4.9% | 0.44 (0.36-0.53)†† | 89.4% | 6.93 (6.12-7.87)†† |

*ED LOS plus Inpatient LOS for patients who did not expire; excludes patients who died within 72 hours

**CNS infection, intraabdominal infection, bacteremia, gangrene, neutropenic fever, orthopedic infection, intrathoracic infection

***Categories of infection have OR relative to pneumonia

****Diagnosis associated with need for surgical intervention or hospitalization

Table S4-A shows univariable odds ratios and 95% confidence intervals for associations between primary emergency department diagnostic categories and two reference outcomes (hospital length of stay <72 hours and no ICU stay)in a cohort of 9,443 patients placed on a sepsis pathway in the emergency department of five hospitals in a major urban health system from 2022-2024.  Categories were modeled on the schema of Linden and Melhammar and assigned manually by the principal investigator using data obtained from an electronic health record query.

Table S4-B: Univariable Associations of Diagnoses with Outcomes of Patients Placed on Sepsis Pathway

| | Survival to discharge | | No sepsis adjudication | |
|---|---|---|---|---|
| | % | OR** | % | OR** |
| All patients | 84.5% | -- | 28.0% | -- |
| Sepsis | 82.2% | 0.51 (0.42-0.62)†† | 7.1% | 0.61 (0.48-0.76)†† |
| Serious Infection*** | 93.2% | 1.51 (1.00-2.38) | 10.1% | 0.90 (0.61-1.28) |
| Infection | 90.0% | reference | 11.2% | Reference |
| Pneumonia | 85.2% | reference*** | 11.7% | reference*** |
| GU infection | 93.6% | 2.55 (1.68-4.00)†† | 9.2% | 0.76 (0.51-1.12) |
| GI infection | 92.8% | 2.24 (1.23-4.49)† | 16.3% | 1.47 (0.90-2.33) |
| SSTI | 97.2% | 6.09 (3.02-14.55)†† | 6.3% | 0.51 (0.28-0.85)† |
| ENT infection | 93.3% | 2.43 (0.48-44.18) | 20.0% | 1.89 (0.43-6.25) |
| Fever | 91.5% | 1.88 (1.03-3.78) | 13.8% | 1.22 (0.69-2.04) |
| Unspecified/other | 96.2% | 4.33 (0.91-77.77) | 13.0% | 2.27 (0.81-5.56) |
| Viral Infection | 85.2% | 0.64 (0.44-0.94)† | 84.0% | 50.0 (33.3-50.0)†† |
| Organ Failure | 75.2% | 0.34 (0.27-0.41)†† | 22.7% | 3.85 (3.23-4.76)†† |
| Non-infectious | 85.9% | 0.67 (0.56-0.81)†† | 47.8% | 7.14 (6.25-8.33)†† |
| Possible infection | 81.2% | 0.48 (0.39-0.59)†† | 34.0% | 4.17 (3.45-5.00)†† |
| Non-severe | 88.7% | reference | 27.5% | reference |
| Severe dx**** | 79.1% | 0.49 (0.43-0.55)†† | 34.2% | 1.37 (1.23-1.52)†† |
| Other | 87.1% | 0.86 (0.73-1.01) | 17.6% | 0.56 (0.50-0.65)†† |
| Boarding (ED LOS >24 hours) | 89.4% | 1.83 (1.61-2.09)†† | 26.9% | 0.93 (0.84-1.02) |

*ED LOS plus Inpatient LOS for patients who did not expire; excludes patients who died within 72 hours

**CNS infection, intraabdominal infection, bacteremia, gangrene, neutropenic fever, orthopedic infection, intrathoracic infection

***Categories of infection have OR relative to pneumonia

****Diagnosis associated with need for surgical intervention or hospitalization

Table S4-B shows univariable odds ratios and 95% confidence intervals for associations between primary emergency department diagnostic categories and two reference outcomes (survival to discharge and not being adjudicated as having sepsis by the healthcare system's institutional sepsis committee) in a cohort of 9,443 patients placed on a sepsis pathway in the emergency department of five hospitals in a major urban health system from 2022-2024. Categories were modeled on the schema of Linden and Melhammar and assigned manually by the principal investigator using data obtained from an electronic health record query.
